# Predictors of second-line antiretroviral treatment virological failure at Felege hiwot and University of Gondar comprehensive specialized hospitals Amhara region, Northwest Ethiopia: a case-control study

**DOI:** 10.1101/2023.07.20.23292946

**Authors:** Getahun Ayenew Wubetu, Yeshambel Agumas Ambelie, Tebkew Shibabaw, Gebremariam Getaneh, Michael Getie Abate

## Abstract

**Background:** Second-line HIV treatment failure has become increasing worldwide, mainly in sub- Sahara Africa including Ethiopia. Even though the problem becomes increasing, inadequate information was available about its magnitude and predictors in the current study area.

**Objective:** To assess the predictors of second line Anti-Retroviral Treatment virological failure among second line ART users.

**Method and materials:** Institutional based unmatched case control study design was conducted from first September 2021 to December last 2021 at Felege Hiowt and University of Gondar Comprehensive Specialized Hospitals; Amhara region, Northwest Ethiopia. A total of 216 patients (60 cases and 156 controls) were recruited by Simple random sampling technique with 1:3 cases-to-controls ratio. Patients who had two viral load results ≥1000 copies/ml within a 3-month interval after taking ART drugs for at least 6 months were cases whereas ≤1,000 copies/ mL were controls. The sample size was calculated by using Epi-Info version 7.2.4. Structured questionnaires were used to gather the required information. SPSS version 26 was used to summarize the findings. In bivariate logistic regression model, Variables with two-tailed P-value ≤ 0.25 at 95% confidence interval were transferred into multivariate binary logistic regression mode and P value at ≤ 0.05 was set as statistically significant.

**Results:** Out of 216 patients recruited, 212 were participated with a response rate of 98.2%. Among the participants, 117 (55.2%) were males and 187 (88.2%) were urban dwellers. 208 (98.1%) of the respondents had age > 24 years, 73 (34.4%) had elementary level of education, 72(34%) had poor ART adherence and 112(52.8) did not disclose their HIV status. Likewise, most of the patients 147(69.37) didn’t used condom. The Predictors were not disclosing HIV status (AOR=3.4, 95% CI: 1.52 – 7.79), poor adherence level (AOR=5.27, 95% CI: 2.2 - 12.5), not using condom (AOR=4.47, 95% CI: 1.63 – 12.2) and high Viral load (≥1000 copes/ml) when switched to second line ART (AOR=3.56, 95% CI: 1.5 - 8).

**Conclusion and recommendations:** The Predictors of second line Anti-Retroviral Treatment virological failure were non-disclosure, poor adherence, not using condom and high Viral load (≥1000 copes/ml) at switched to second line ART. Disclosing their HIV status, using condom and improving their adherence level for patients and counselling about the importance of disclosure and good adherence for health care providers are crucial.

## INTRODUCTION

Human immunodeficiency virus (HIV) is a single-stranded RNA virus which attacks the body’s immune system, specifically the white blood cells called CD4 cells. HIV has been continued to be a major global public health issue, having claimed almost 36.3 million lives so far. About 79.3 million people have become infected with HIV since the start of the epidemic. Only in 2020, there were 37.7 million people (1.7 million children <14 years) living with HIV, 1.5 million People became newly infected and 680 000 people died from AIDS-related illnesses worldwide. However, with increasing access to effective HIV prevention, diagnosis, treatment and care including for opportunistic infections, HIV infection has become a manageable chronic health condition enabling people living with HIV to lead long and healthy lives (1,2)

But the emergence of drug-resistant HIV has the potential to become a major public health threat worldwide as it limits treatment options for people living with HIV. HIV drug resistance (HIVDR) is an alteration in the genetic structure of HIV that affects the ability of a specific drug or combination of drugs to block replication of the virus. HIVDR occurs when the virus starts to make changes (mutations) to its genetic make-up (RNA) that are resistant to certain HIV drugs, or classes of HIV drugs. Now a day’s, global prevalence of HIVDR is rising mainly due to resistance to non-nucleoside reverse transcriptase inhibitor (NNRTI) drugs which make up the backbone of first-line antiretroviral treatment regimes. It is also a serious emerging threat to the global scale-up of HIV treatment access, viral load testing access, monitoring of other clinical factors relating to patient care, patient behavior and clinic and program management (3).

Second-line HIV treatment failure has also become highly prevalent in Sub-Saharan Africa (SSA) with alarming rates during the 12–18 month period of treatment start in children (4). Second-line ART regimens are used when patients develop treatment failure for the first-line treatment regimens (5). If a person on ART who has taken ART drugs for at least 6 months has two consecutive viral load measurements greater than 1000 copies/ml within a 3-month interval with adherence support between measurements, the results will confirm failure of the current treatment regimen and the client needs to be switched to appropriate third line regimen (6,7).

Viral load failure is used as a golden approach to confirm treatment failure. For HIV patients who start ART, Viral load testing should be performed early after initiating ART within 6 months, at 12 months and then at least every 12 months to detect treatment failure. In settings where routine viral load monitoring is available, CD4 cell count monitoring can be stopped in individuals who are stable on ART and virally suppressed (6,8).

The rise in Antimicrobial Resistance (AMR) is one of the greatest threats to global health. If it is not urgently addressed, it may result in millions of deaths, an increase in new and hard-to-treat infections and increased health-care costs. After 15 years of global scale-up of antiretroviral therapy (ART), rising prevalence of HIV drug resistance in many low and middle income countries poses a growing threat to the HIV response with an increase in mortality, morbidity, HIV incidence and health care costs (3,8). All current antiretroviral drugs (ARVD), including newer classes, are at risk of becoming partly or fully inactive because of the emergence of drug-resistant virus. People receiving ART can acquire HIVDR or people can also be infected with HIV that is already drug resistant (9). The issue is even more problematic in prevention of mother-to-child HIV transmission (PMTCT) in HIV programs (10).

Globally, the prevalence of DRHIV to NNRTI drugs has significantly increased since 2001. This rise has been observed more rapidly in Eastern Africa with estimated annual incremental increase of 29%, Southern Africa (23%), Western and Central Africa (17%), Latin America (15%) and Asia (11%). In eastern Africa, the Prevalence of pretreatment NNRTI resistance increases from 0% in 1996 to 13% in 2016 (3,11).

Virological ART failure is highly prevalent in sub-Saharan Africa (10). Especially in low-income countries, HIV drug resistance is still a serious threat to their health (12). Among populations receiving NNRTI-based ART with viral non-suppression, the levels of NNRTI and NRTI resistance ranged from 50% to 97% and from 21% to 91% respectively. Estimates of dual class resistance (NNRTI and NRTI) ranged between 21% and 91% of individuals for whom NNRTI-based first-line ART failed (13).

A survey implemented between 2014 and 2018 in 39 countries, showed that Pretreatment HIV Drug Resistance (PDR) to efavirenz/nevirapine among adults starting or restarting first-line ART ranges 10.2% in NEPAL to 25.9% in Honduras. Pretreatment drug resistance among treatment-naive infants newly diagnosed with HIV from 2012 to 2018 was very high, ranging from 34% in Swaziland to 69% in Malawi. Overall, the prevalence of any HIVDR among all individuals receiving treatment between 2014 and 2018 were ranged from 3% in Viet Nam to 29% in Honduras (13).

In Sub Sahara Africa (SSA), Second-line HIV treatment failure has become highly prevalent with alarming rates during the 12–18 month period of treatment start (4). Among patients receiving protease inhibitor (PI) based 2nd-line HIV treatment, 25% experienced virological failure at some point during follow-up in this region in 2016 (14). Another report in the same year in Sub-Saharan Africa showed that second line ART drug resistance mutation were 14% (15). A systematic review and meta-analysis conducted in SSA in 2019 also reported that, Second-line HIV treatment failure was 13.4%. Of which 65% PI-based and 35% was Ritonavir boosted PI-based second-line ART regimens (4).

In resource-limited settings, the prevalence of second-line virological ART failure rates among adults in 2012 were 21.8–33%, 14–38%, 15—38.5% and 10–38.0% at 6, 12, 24, and 36 months of initiation on second-line ART respectively (16). There were 1668 patients with virological failure in south Africa in 2012 (17), 109 patients with confirmed virological failure in Malawi in 2010 (18) and Second-line ART Virological failure was 18% in Rwanda by the end of 2016 (19). Some Studies in Tanzania and southeast Uganda also indicated that there was a high incidence of virological failure (20).

The number of virological treatment failure is increasing from time to time in Ethiopia (21). It becomes a major challenge for HIV ADIS prevention and control mechanisms (22). The adjusted magnitude of VF among population taking ART in Ethiopia was found to be 11% (23). According to a study done among children and adolescents on second line ART in a pediatric cohort of an Ethiopian tertiary hospital, proportion of treatment failure (TF) were 14 out of 76 patients(24). A high incidence rate of second-line treatment failure (9.86 per 100 person-years or 254/1011) was also noticed in Amhara Region in 2016 (5).

Among the determinants of second line ART failure, poor adherence to ART, high viral load, not disclosure about HIV status, opportunistic infection, low CD4 counts < 350 cell/mm^3^, low BMI (< 16 kg/m^2^), young age 15– 29 year patients, smoking and drug abuse were the main factors (5,25).

Even if the problem is growing, there is still a paucity of information in developing countries on the factors for second-line ART failure (18). Identifying risk factors of second-line treatment failure remains crucial to reduce the need for third-line drugs (26). Even though the incidence and prevalence of second line ART failure becomes increasing, inadequate information is available about the magnitude and its predictors of second line ART virological failure in the current study area. Most of the previous studies were focused on adults >18 years. Therefore, identifying the Predictors of second-line antiretroviral treatment virological failure among patients on antiretroviral therapy was the main objective of this study.

Findings of this research will give information about the risk factors of second line virological ART failure in the study area. It will serve as a base line data for treatment centers to address factors related to second line treatment failure. The result might be useful for stakholders to set programs and action plans on the need to third line regimens. It will be used as a source of information for researchers. It will also give important recommendations for patients on their treatment.

## METHODS AND MATERIALS

### Study settings

The study was conducted at Felege hiwot and University of Gondar comprehensive specialized hospitals Amhara region, Northwest Ethiopia. FHCSH is found in Bahir Dar city, the capital city of Amhara Regional state. It is located 561km away from Addis Abeba. Geographical coordinates of Bahir Dar city are 11° 36’ 0” North, 37° 23’ 0” East and an Elevation of 1,800 m (5,900 ft.) above sea level (52,53). According to the Amhara National Regional State Plan Commission Bureau 2021 data, Bahir Dar city administration had an estimated total population of 406,433 (192,041 male and 214,393 female) dwellers. Of which 85% live in the urban and 15% in the peri-urban and rural areas of the city. Based on Amhara Regional health bureau 2021 report, 14 Health posts, 10 Health centers 2 general hospitals and 2 referral hospitals are governmental health institutions found in the city. There are also 10 basic clinics, 28 Medium clinics, 10 higher clinics and 5 general hospitals among private health institutions. Only one governmental specialized referral hospital (Felege Hiwot) has provided third line ART treatment service in Bahir Dar. There were 740 and 31 ART patients who were on second line and third-line regimens respectively at FHCSH by the end of August 2021.

The city of Gondar is situated in North-western parts of Ethiopia, Amhara Regional State. It far 725 km from Addis Ababa, 175 km from Bahir Dar and 120km from the Simien Mountains. It founds at 12° 36′ 0″ N, latitude 37° 28′ 0″ E longitude coordinates and an elevation of 2133 m above sea level. (54,55). The Regional Plan Commission Bureau also reported that, the metro area population of Gondar city in 2021 was 454,445 (218,378 male and 236,068 female). UGCSH is used as the referral center for more than 7 million catchment population (56). Based on reports obtained from this hospital, there were 5513 patients on ART until September 1^st^, 2021. Out of these 520 were second-line ART users and 29 were on third-line regimens.

### Study design and period

Institutional based unmatched case control study design was conducted from September first, 2021, to December last 2021.

### population

All HIV patients at FHCSH and UGCSH who were on second line and third line ART regimens and patients who were on second line and third line ART regimens and took second line ART at least for six months were the Source population and Study population respectively.

### Inclusion and exclusion criteria

All HIV patients who were on second line and third line ART regimens at FHCSH and UGCSH ART clinics during the study period were included in the study. Whereas those patients who had not taken second line ART regimen for more than six months during the study period were excluded in this study.

### Sample size determination and sampling techniques

Sample size was calculated using EPIINFO version 7.2.4 (a two-population proportion formula). n= sample size (number of participants), 95% confidence level) Z α/2=1.96 (for 0.05 significance level), for 80% power, Z_β_= 0.84, r = 1/3 (the ratio of cases to controls), p1= 31% (% of cases among exposed), p2= 11.2% (% of cases among non-exposed). From previous study conducted in Wollo, Amhara Regional State, Northeast Ethiopia, the following significant predictors for second-line antiretroviral treatment virological failure were found, Age 15 – 29 years, Poor adherence, Have no disclosure status, BMI < 16 k.g/m^2,^ CD4 count < 100cells/mm^3^, CD4 count 100 – 350 cells/mm^3,^ Having opportunistic infections (51). We used this report as a base line data, the minimum sample size including 10% non-respondent rate for this study was 206 participants (52 cases and 154 controls). But we recruited 216 patients (all the cases 60) and 156 controls.

### Sampling techniques

#### Case selection

All HIV patients who had taken a second line ART for at least six months and had developed a virological failure were selected and listed from the patients registration book at FHCSH and UGCSH during the study period. There were 31 patients at FHCSH and 29 patients at UGCSH who were failed for second line ART. **Control selection:** All HIV patients who had taken a second line ART regime at least for six months and didn’t developed virological ART failure were selected and listed from the patients registration book at FHCSH and UGCSH during the study period. Participants were recruited by a simple random sampling technique (generated using a computer excel).

By considering proportional allocation method,

Let nf = number of control participants and Nf = total number of controls from FHCSH and ng = number of control participants and Ng = total number of controls from UGCSH,

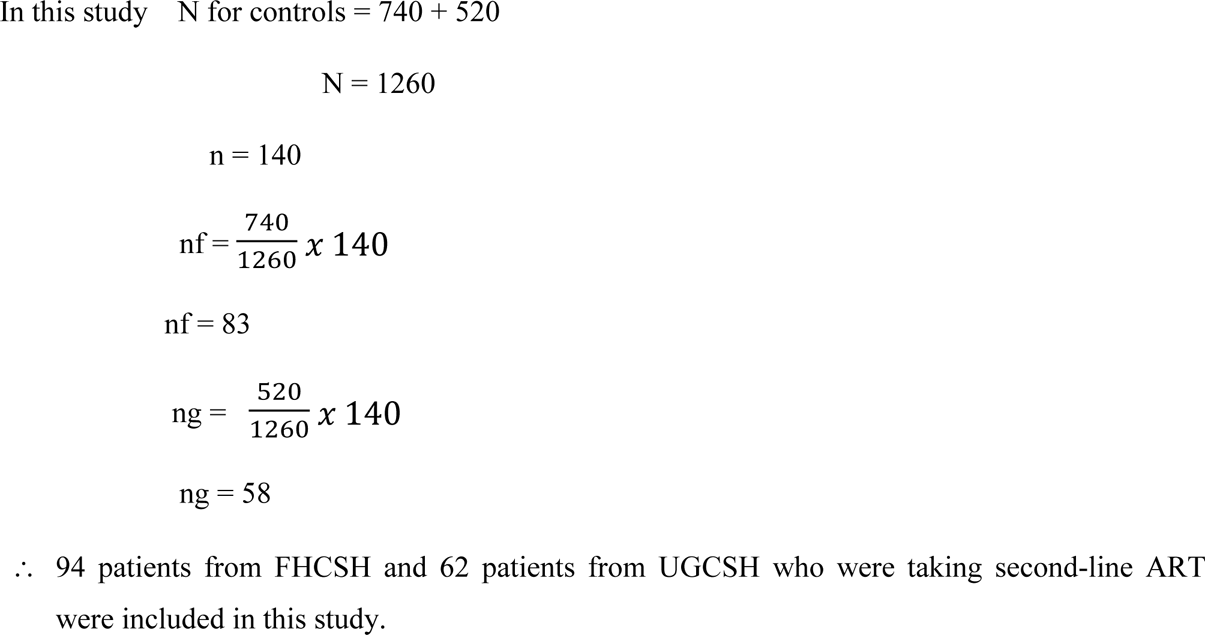

### Definition of terms

**Stable on ART-** HIV Patients on ART for at least 1 year, no current illnesses or pregnancy, good understanding of lifelong adherence and evidence of treatment success (two consecutive viral load measurements below 1000 copies/mL) and absence of adverse drug reactions requiring regular monitoring (57).

**Adherence**- Refers to the whole process from starting HIV treatment, keeping all medical appointments and taking HIV medicines every day and exactly as prescribed. Or it is the degree to which the person’s behavior, taking medication, following a diet and/or changing lifestyle corresponds with the agreed recommendations from a health care provider (33).

**Good Adherence;** drug adherence of ≥95% or if <2 doses of 30 doses or <3 doss of 60 doses is missed as documented by ART healthcare provider (33).

**Fair/medium/ adherence**. Drug adherence of 85–94% or 3–5 missed drug doses out of 30 doses or 4–9 missed drug doses out of 60 doses (33).

**Poor adherence;** If drug adherence is <85% or ≥6 doses of missed ART drug doses out of 30 doses or >9 doses of missed ART drug doses out of 60 as documented by the ART healthcare provider (33).

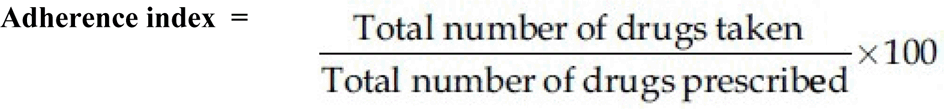

**Suboptimal adherence**; a report of ≥1 reason for missing ART ≥5 times within the past month (58).

**Opportunistic Infections (OI);** Infections that develop as a result of HIV-inflicted damage to the immune system (TB, PCP, gastrointestinal OI, herpes simplex, herpes zoster, fungal infection) and other national ART guidelines define opportunistic infection (59)

**Virological failure;** refers to a persistently detectable viral load exceeding 1000 copies/mL (two consecutive viral load measurements within a 3-month interval with adherence support between measurements) after at least 6 months of using ART (8)

**Non-Disclosure:** patients who did not disclose about their HIV status for anyone except their couples and health care providers.

### Study Variables

Independent variables were Socio demographic, behavioral, clinical, and immunological characteristics of the patient. Of these; sex, age, weight, residence, marital status, educational status, job, alcohol intake, drug abuse, smoking, disclosure status, number of sexual partners, condom use, adherence, body mass index (BMI) at switch, opportunistic infections (OI), TB coinfection, nutritional status, WHO HIV clinical staging at switch to second line, CD4 count (CD4 count at 2^nd^ line ART initiation), viral load, type of first and second line treatment (being on a protease inhibitor plus 2 nucleoside analogues(NRTIs) or not), duration in second line ART and duration in first line ART. Whereas the dependent variable (outcome of interest) was second line ART virological failure.

### Data collection tools and techniques

The data were collected by using structured questionnaires and checklist prepared by the principal investigator. The tools were adapted from different literatures, WHO guidelines, institutional checklists, and some of it were developed by the investigator. This tool includes socio demographic, behavioral, clinical, and immunological related data. Personal interview and patient registration book/ card/ review was performed using data collection tools to gather the required information.

### Data quality managements

To maintain the data quality, different techniques were employed to address major source of errors. The questionnaires were translated to local language (Amharic). Health professionals who were participated in the data collection were got on sight training before the data collection. Each day the collected data were crosschecked for its completeness, accuracy, clarity, consistency and missed values and missed variables carefully. Necessary modifications were made based on the gaps identified. All collected data were entered into EPI-data version 3.1 after being coded. Then it was transferred to Statistical Package for Social Science (SPSS) version 26 for analysis.

### Data processing and analysis

The collected data were entered and analyzed by using SPSS version 26 for windows. Descriptive statistics was used to present and summarize the findings. Bivariate and multivariate binary logistic regression model was employed to identify determinates of second line ART virological failure. Odds ratios with its 95% confidence intervals and two-tailed P-value was calculated. Variables with P-value ≤ 0.25 in the bivariate analysis were included in the multivariate logistic regression mode and variables which had P value ≤ 0.05 in the multivariate analysis were consider as statistically significant.

### Ethical clearance

Ethical clearance was obtained from Bahir Dar University, College of Medicine, and Health Sciences Institutional Review Board (IRB). Official letter of co-operations was provided to FHCSH and UGCSH prior to data collection. Assent from children and verbal informed consent from adults were obtained after explaining the purpose and objective of the study. Confidentiality of the data had been maintained.

## RESULTS

### Socio-demographic and Behavioral characteristics

Out of the total of 216 patients on second- and third-line ART (60 cases and 156 controls) recruited, 212 (59 cases and 153 controls) were participated in this study. This accounts for a response rate of 98.2%. The mean age of the participants were 40.7 years. Among the participants, 117 (55.2%) were males and 187 (88.2%) respondents were urban residents. 208 (98.1%) of the respondents had age > 24 years and 4 (1.9%) were 24 years or below. 102 (48.1%) were married participants and 73 (34.4%) had elementary level of education (Table 1).

**Table 1:** Socio demographic characteristics of patients on second- and third-line ART regimen at FHCSH and UGCSH; Amhara Region, Northwest Ethiopia from September to December 2021.

| General variables | Variables category | Frequency of Virological failure. |  | Total No. (%) |
| --- | --- | --- | --- | --- |
|  |  | No. (%) | No. (%) |  |
|  |  | Cases (N=59)<br>No. (%) | Controls (N=153)<br>No. (%) | N = 212 |
| Sex | Male | 33(55.9) | 84(54.9) | 117(55.2) |
|  | Female | 26(44.1) | 69(45.1) | 95(44.8) |
| Age (in year) | < 14 | 3(5.1) | 1 (0.7) | 4(1.9) |
|  | 14 – 24 | 4(6.8) | 13(8.5) | 17(8.1) |
|  | >24 – 64 | 51(86.4) | 136(88.9) | 187(88.2) |
|  | >64 | 1(1.8) | 3(2) | 4(1.9) |
| Residence | Urban | 51(86.4) | 136(88.9) | 187(88.2) |
|  | Rural | 8(13.6) | 17(11.1) | 25(11.8) |
| Educational status | Illiterate | 14(23.7) | 43(28.1) | 57(26.9) |
|  | Elementary level | 21(35.6) | 52(34) | 73(34.4) |
|  | Secondary level | 12(20.3) | 21(13.7) | 33(15.6) |
|  | Preparatory level | 11(18.6) | 20(13.1) | 31(14.6) |
|  | College and above | 1(1.7) | 17(10) | 16(7.5) |
| Marital status | Single | 16(27.1) | 48(31.4) | 64(30.2) |
|  | Married | 34(57.6) | 68(44.4) | 102(48.1) |
|  | Divorced | 7(11.9) | 25(16.3) | 32(15.1) |
|  | Widowed | 2(3.4) | 12(7.8) | 14(6.6) |

From the study participants, 33 (55.9%) of the cases and 3(2%) of the controls had poor adherence to ART treatments. Similarly, 46 (78%) of the cases and 66 (43.1%) of the controls did not disclose their HIV status during taking ART. Likewise, 9 (15.3%) of the cases and 26 (17%) of the controls had a history of alcohol use. Most of the cases 53 (89.8%) and the controls 94 (61.4%) did not used condom (Table 2).

**Table 2:** Behavioral characteristics of patients on second- and third-line ART regimen at FHCSH and UGCSH; Amhara Region, Northwest Ethiopia from September to December 2021.

| General variables | Variables category | Frequency of Virological failure.<br>No. (%) |  | Total No. (%) |
| --- | --- | --- | --- | --- |
|  |  | Cases (N=59)<br>No. (%) | Controls (N=153)<br>No. (%) | 212 |
| Alcohol consumption | Yes | 9(15.3) | 26(17) | 35(16.5) |
|  | No | 50(84.7) | 127(83) | 177(83.5) |
| Using condom | Yes | 6(10.2) | 59(38.6) | 65(30.7) |
|  | No | 53(89.8) | 94(61.4) | 147(69.374.5) |
| Disclosure status | Disclosed | 13(22) | 87(56.9) | 100(47.2) |
|  | Not disclosed | 46(78) | 66(43.1) | 112(52.8) |
| Smoking cigarettes | Yes | 5(8.5) | 1(0.7) | 6(2.8) |
|  | No | 54(91.5) | 152(99.3) | 206(97.2) |
| Level of Adherence | Good | 13(22) | 91(59.5) | 104(49) |
|  | Medium | 13(22) | 39(25.5) | 36(17) |
|  | Poor | 33(55.9) | 3(2) | 72(34) |

### Clinical and immunological characteristics

Among the participants recruited, 86(40.6%) had a CD4 count <200 (cells/mm3). 8(13.6%) of the cases and 7(4.6%) of the controls were severely malnourished. Viral load when switched to second line ART is ≥1000(copes/ml) for 71(33.5) patients. Most of the participants were at WHO HIV clinical stage I (Table 3).

**Table 3:** Clinical and immunological characteristics of patients on second- and third-line ART regimen at FHCSH and UGCSH; Amhara Region, Northwest Ethiopia from September to December 2021.

| General variables | Variables category | Frequency of Virological failure. |  | Total No.<br>(%)<br>N = 212 |
| --- | --- | --- | --- | --- |
|  |  | Cases (N=59)<br>No. (%) | Controls (N=153)<br>No. (%) |  |
| Body mass index<br>(BMI) | <18.5 | 20(33.9) | 25(16.3) | 45(21.2) |
|  | 18.5 – 25 | 36(61) | 106(69.3) | 142(67) |
|  | >25 – 30 | 2(3.4) | 21(13.7) | 23(10.8) |
|  | >30 | 1(1.7) | 1(0.7) | 2(0.9) |
| CD4 count<br>(cells/mm3) | <200 | 24(40.7) | 62(40.5) | 86(40.6) |
|  | 200 – 350 | 21(35.6) | 53(34.6) | 74(34.9) |
|  | 350.01 - 500 | 9(15.3) | 29(19) | 38(17.9) |
|  | >500 | 5(8.5) | 9(5.9) | 14(6.6) |
| Nutritional status | Normal | 33(55.9) | 104(68) | 137(64.6) |
|  | MAM (Moderately) | 10(16.9) | 25(16.3) | 35(16.5) |
|  | SAM (severely) | 8(13.6) | 7(4.6) | 15(7.1) |
|  | Over weight | 8(13.6) | 17(11.1) | 25(11.8) |
| Co-trimoxazole<br>started | Yes | 28(47.5) | 74(48.4) | 102(48.1) |
|  | No | 31(52.5) | 79(51.6) | 110(51.9) |
| Fluconazole<br>started | Yes | 3(5.1) | 22(14.4) | 25(11.8) |
|  | No | 56(94.9) | 131(85.6) | 187(88.2) |
| Viral load when<br>switched to second<br>line ART (copies/ml) | <150 | 18(30.5) | 104(68) | 122(57.5) |
|  | 150 – 999.999 | 11(18.6) | 8(5.2) | 19(9) |
|  | ≥1000 | 30(50.8) | 41(26.8) | 71(33.5) |
| WHO HIV clinical<br>stage | stage I | 49(83.1) | 151(98.7) | 200(94.3) |
|  | stage II | 1(1.7) | 1(0.7) | 2(0.9) |
|  | stage III | 9(15.3) | 1(0.7) | 10(4.7) |
| Duration in first line<br>ART /in years/ | <2 years | 6(10.2) | 26(17) | 32(15.1) |
|  | 2 -5 years | 16(27.1) | 58(37.9) | 74(34.9) |
|  | 5.01 – 10 years | 30(50.8) | 60(39.2) | 90(42.5) |
|  | > 10 years | 7(11.9) | 9(5.9) | 16(7.5) |

### Factors associated to second line ART virological failure

In bi-variable logistic regression from a total of 36 variables the following: Not using condom, Non-disclosure about HIV status, poor Level of Adherence, severely malnutrition nutritional status and Viral load ≥1000 copes/ml when switched to second line ART were significantly associated with second line virological ART failure (P-value<0.25). These factors were also selected for further multivariable binary logistic regression analysis (Table 4).

**Table 4:** Factors related to second line ART Virological failure among HIV Patients at FHCSH and UGCSH; Amhara Region, Northwest Ethiopia from September first, 2021, to December last 2021.

| General variables | Variables category | Virological failure |  | COR (95% CL) | AOR (95% CL) |
| --- | --- | --- | --- | --- | --- |
|  |  | Yes | No |  |  |
| Using condom | Yes | 6 | 59 | 1 |  |
|  | No | 53 | 94 | 5.54 (2.3 - 13.7)**** | 4.5(1.63 – 12)*** |
| Disclosure status | Disclosed | 13 | 87 | 1 |  |
|  | Not disclosed | 46 | 66 | 4.6(2.33 – 9.34)**** | 3.4(1.5 – 7.8)*** |
| Level of Adherence | Good | 13 | 91 | 1 |  |
|  | Medium (faire) | 13 | 23 | 3.95(1.6 - 9.7)*** | 3.7(1.3 - 10.7)* |
|  | Poor | 33 | 39 | 5.9(2.8 – 12.4)**** | 5.3(2.2 – 12.5)**** |
| Nutritional status | Normal | 33 | 104 | 1 |  |
|  | Moderately malnourished | 10 | 25 | 1.26(0.55 – 2.9) | 1.2(0.4 – 3) |
|  | Severely malnourished | 8 | 7 | 3.6(1.2 – 10.7)* | 3.56(1 – 14.4) |
|  | Over weight | 8 | 17 | 1.48(0.59 – 3.8)* | 1.6 (0.4 – 5) |
| Viral load when switched to second line ART (copies/ml) | <150 | 18 | 104 | 1 |  |
|  | 150 – 999.9 | 11 | 8 | 7.9(2.8 – 22)**** | 5.4(1.5 – 19)** |
| | $\geq 1000$ | 30 | 41 | 4.2(2.13 – 8.4)**** | 3.56(1.5 - 8)*** |

Note; P value $< 0.05$ = \*, P value $< 0.01$ = \*\*, P value $< 0.005$ = \*\*\*, P value $< 0.001$ = \*\*\*\*

In the multivariable binary logistic regression analysis, Not using condom, non-disclosure about HIV status, poor level of adherence and viral load ≥1000 copes/ml when switched to second line ART had a significant impact on second line ART outcomes. Peoples who did not disclosed their HIV status were 3 times (AOR=3.4, 95% CI: 1.52 – 7.79) more likely to develop second line ART virological failure as compared to those patients who disclosed their HIV status. Among the respondents those patients who had poor level of adherence were 5 times (AOR= 5.27, 95% CI=2.2 – 12.5) more likely to develop second line ART virological failure as compared to those who had a good adherence (**Error! Reference source not found.**).

Moreover, the likely hood of developing second line virologic failure among patients with Viral load ≥1000 copes/ml when switched to second line ART medication were 3.5 times (AOR=3.56, 95% CI: 1.5 - 8) more likely as compared to those patients who had Viral load result <1000 copes/ml (**Error! Reference source not found.**).

## Discussion

This study was aimed to assess the predictors of virological failure among second-line ART users. Poor adherence to ART, non-disclosure of HIV status, not using condom and having Viral load ≥1000 copes/ml when switched to second line ART were the factors which increased the odds of second line ART virological failure.

Patients who had poor ART adherence were 5 times more likely to develop second line ART virological failure as compared to patients who had good treatment adherence. Similar findings was reported in Wollo, Amhara Regional State Northeast Ethiopia (50) and Northern Ethiopia (25) where the odds of developing second line ART virologic failure was strongly associated with poor ART adherence. This finding is supported by a study conducted in Johannesburg, South Africa (26), Southwestern Nigeria (47), sub-Saharan Africa (44) and Asia (42) which showed that poor adherence was a strong predictor of second-line ART failure. Another investigation done in Cambodia (Southeast Asian) also confirmed that, patients on second line ART with good adherence were significantly associated with viral suppression (43).

ART adherence is generally regarded as an important factor in achieving optimal treatment outcomes. High levels of treatment adherence to ART results to effective viral suppression outcomes. Poor adherence to ART is the most frequent cause of treatment failure and the subsequent development of drug resistant strains of HIV. If the patients do not always take their meds on schedule as prescribed the virus will replicate and develop new resistance mutations over time. Missing doses can lead to low drug levels in the body, which allows the virus to resume replication and accumulate resistance mutations as it multiplies. This results less effective viral suppression and a rise in plasma viral load. Inadequate suppression of viral replication has a continued destruction of CD4 cells, progressive decline in immune function and disease progression, Limited future treatment options and higher costs to the individual and ARV program. Even though these risks the immediate health of the patient, it is also an important reason for the emergence of viral resistance to one or more antiretroviral medications (61–63).

The study found that, not using condom was one of the factors for second line ART virological failure. Patients who didn’t used condom were 4 times more likely to develop ART failure than those who used.

Inconsistent use of a condom by people living with HIV/AIDS on ART has led to further risk of new HIV infection and the development of reinfection with new drug-resistant viral strains. Using condom prevents from acquiring and transmitting drug resistant HIV infection. Patients on ART may also acquire transmitted drug resistant HIV virus if they do not use condoms. This type of HIV infection results treatment failure among ARV drug–naive people with no history of ARV drug exposure (34,64). Not using Condom also exposed for many sexually transmitted infections (STIs) that leads to further worsening the HIV infection by dipping the immune system. So that ART may not be effective and new drug resistant viral strains may occurred (65).

The study also identified that patients who had a high viral load (≥1000 copes/ml) when switched to second line ART were 3.5 times more likely to develop second line ART virological failure than those patients who had suppressed viral load (<1000 copes/ml). This outcome is supported by a studies conducted in Sub-Saharan Africa (26) and Johannesburg, South Africa (44) in which they presented that high viral load was a significant predictor for second line ART virological failure.

Different literature stated that a high viral load can lead to a low CD4 cell count which in turn increases the risk of developing an illness or infection. Circulating high viral particles in blood results high destruction of the immune system (CD4 cell) which in turn aggravates disease progression. Finally it will lead to the emergence of resistant viral strains. This indicates the treatment is not working well. The higher the viral load, the more risk developing treatment failure (66).

In addition, our investigation identified that the odds of developing second line virological ART failure was increased by 3 folds for patients who did not disclosed their HIV status as compared to their counter parts.

Previous studies conducted in Wollo, Amhara Regional State, Northeast Ethiopia and Johannesburg, South Africa also reported that not disclosing HIV status was the main risk factors for virological ART failure (50,67).

Different scholars had agreed that disclosure play a significant roles in good ART outcomes (68). It is regarded as a double-edged sword in terms of ART adherence and patient retention in care. People who achieved full disclosure about their HIV status had lower viral load (improved viral suppression), increased ART adherence and allowed for better self-care and treatment. Patients who disclosed about their HIV status have good adherence to ART compared with those who did not. Studies have found that patients who disclosed their sero-status had better social support; stronger family and relationship cohesion; reductions in anxiety and depression; improvements in physical health, emotional support, and financial support; and were better able to take their ART freely. These helps to improve their ART adherence and having lower risk of developing ART failure. Patients who did not disclosed their HIV status were at high risk of developing Virological failure.(69–71).

## conclusions

The study identified that poor adherence to ART medication, not disclosed about HIV status, not using condom and high viral load >1000 copes/ml were significantly associated with second line ART virological failure.

## Recommendations

Patients on ART should not miss their medication for any reasons and use their full efforts to have good ART adherence. Disclosing their HIV status and using condom will have an incredible benefit in terms of social supports and to get early ART treatment. Early identification of patients who had poor adherence, non- disclosed and high viral load will be an important task to reduce ART failure. Such patients will need closer clinical follow-up to minimize the risk of treatment failure. Further investigation is needed to explore factors associated with poor adherence and not disclosing HIV status.

## Data Availability

All relevant data are within the manuscript and its Supporting Information files

## ACKNOWLEDGEMENTS

First, I would like to acknowledge Bahir Dar University College of medicine and health sciences, school of public health, department of health system management and health economics for give me a chance to conduct a research on this topic.

I also acknowledge the Felege Hiwot Comprehensive Specialized Hospital and University of Gondar Comprehensive Specialized Hospital for their co-operative and supportive willingness to my work.

Dr. Yeshambel Agumas and Mr. Tebkew Shibabaw were my advisors whom my gratitude is forwarded for their timely communications and directions on subjects of thesis proposal preparations and wrote this paper.

I would like to thank Mr Melkamu Teshager and Mis Aynalem Mihiret who were participated in my work by data collection.

## ACRONYMS AND ABBREVIATIONS

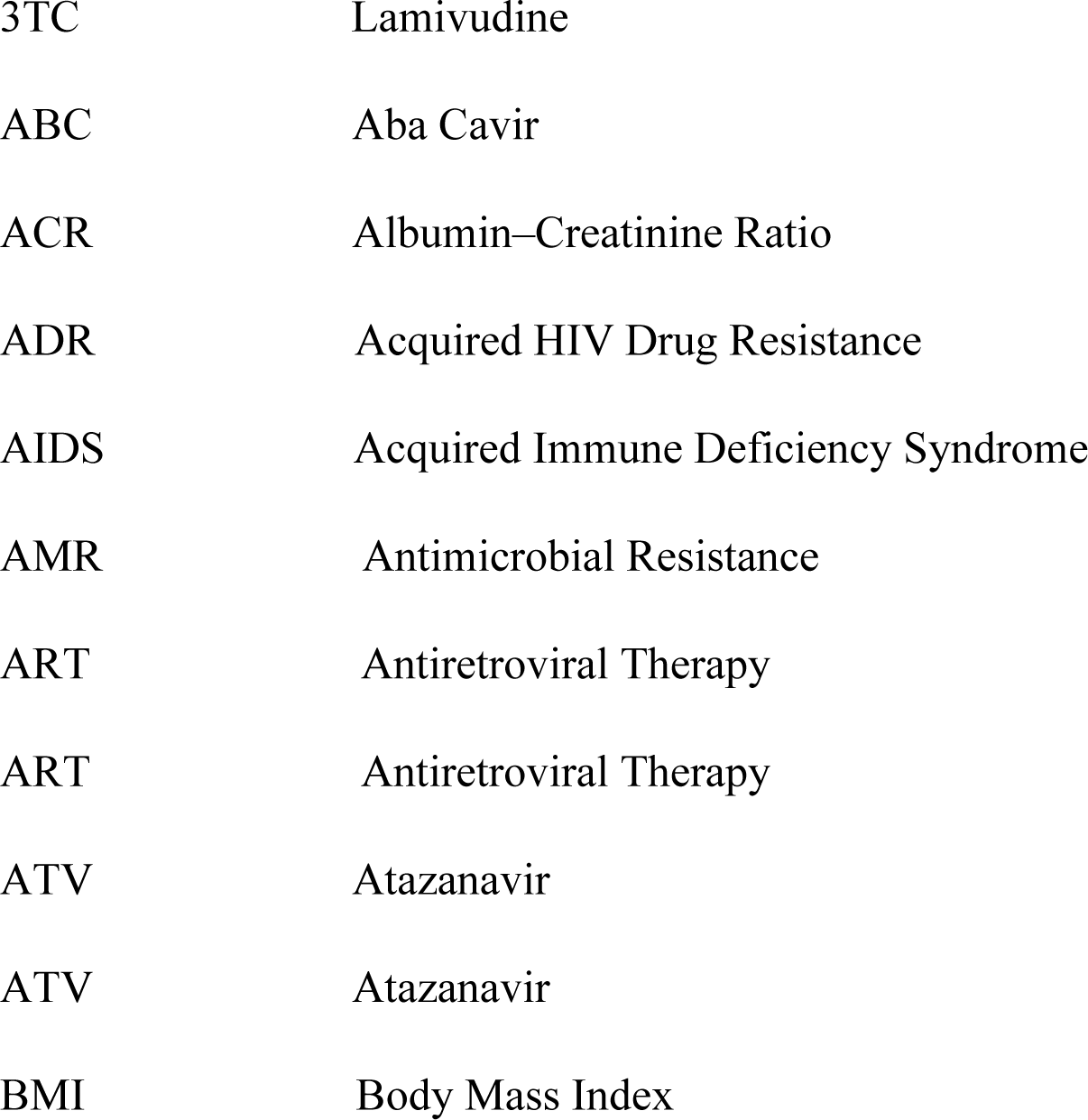
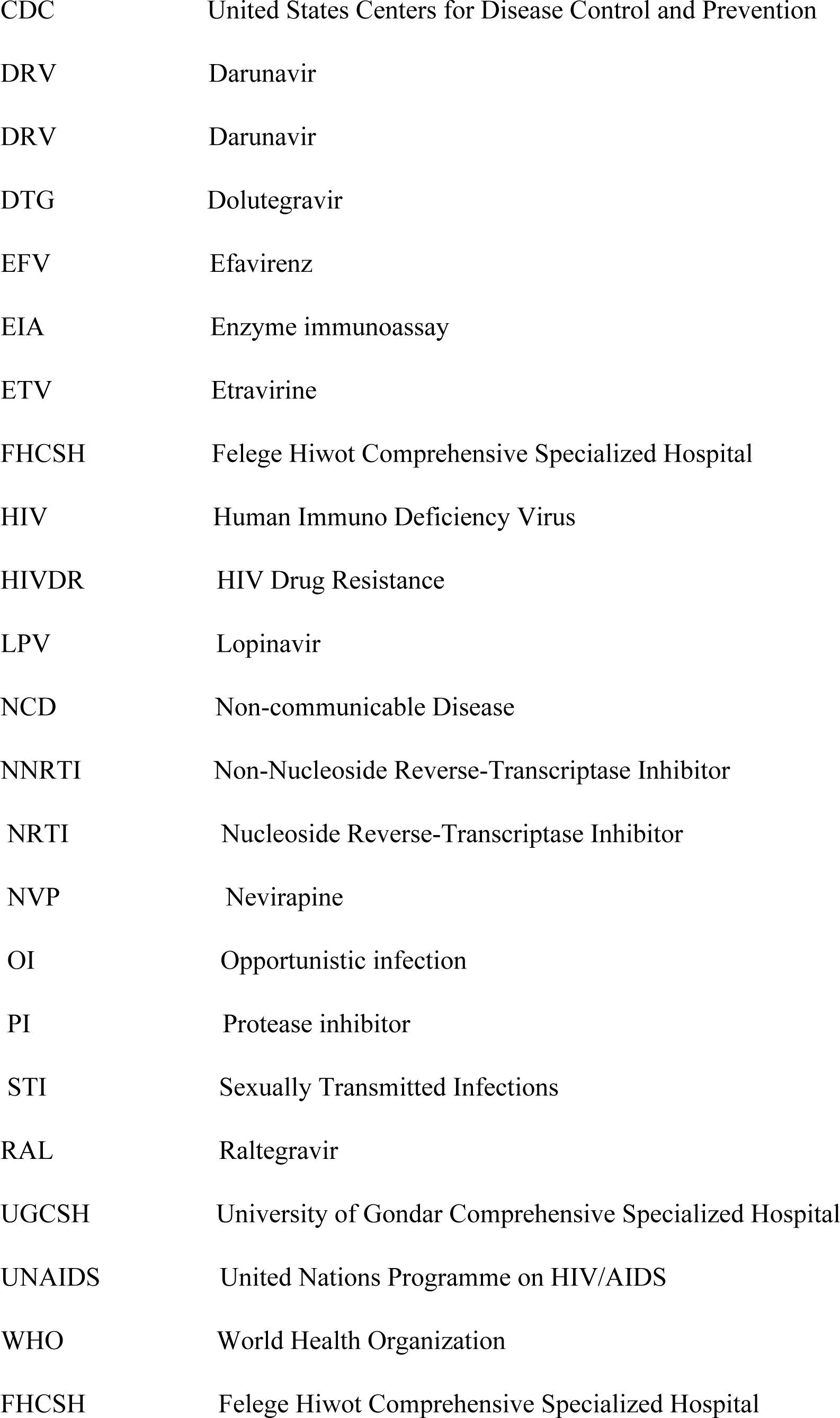
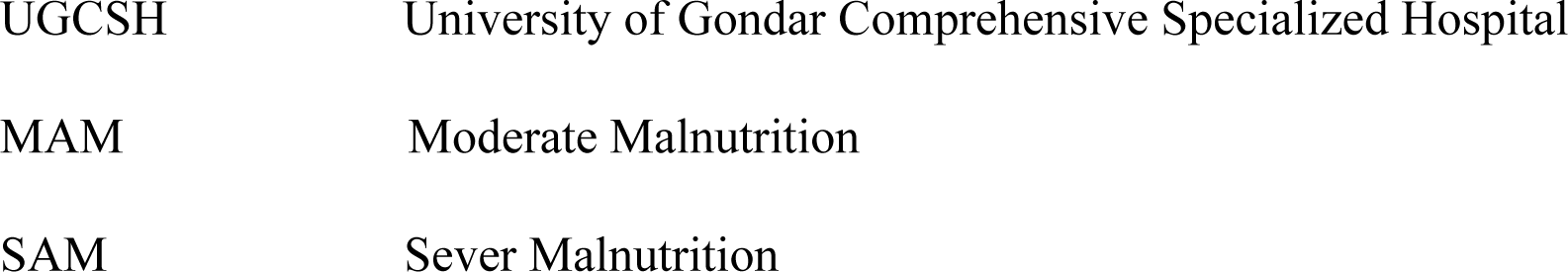

